# Spatial Distribution of Leprosy in Urban Areas: Systematic Review of Environmental and Social Determinants (2015–2024)

**DOI:** 10.1101/2025.09.23.25336438

**Authors:** José Sérgio Macedo Coelho, Thais Ranielle Souza de Oliveira, Diana Leite Alves, Lucas Arruda Macedo Coelho, Carlos Tomaz

## Abstract

Leprosy remains a relevant public health problem in Brazil and other endemic countries, reflecting deep social and environmental inequalities. This systematic review, conducted according to PRISMA 2020 guidelines (PROSPERO registration CRD420251129311), analyzed studies published between 2015 and 2024 that investigated the spatial distribution of leprosy in urban areas. A total of 44 studies were included, mostly conducted in Brazil, complemented by research in India, Indonesia, Bangladesh, Madagascar, and Nigeria. Most studies adopted ecological design and applied techniques such as Moran’s I, SaTScan, Bayesian estimators, Kernel density, and spatial regressions. Results revealed persistent clusters in urban peripheries, particularly in Northern and Northeastern Brazil, as well as a consistent association between leprosy and low income, low education, social inequality, and poor sanitation. International studies confirmed similar patterns in contexts of urban poverty. Methodological appraisal showed that 88.6% of the studies were of high quality. In conclusion, leprosy remains strongly associated with socio-environmental determinants, and spatial analysis is a strategic tool for surveillance and the targeting of public health interventions. This study reinforces the relevance of actions aligned with the Sustainable Development Goals (SDGs), particularly SDG 3 (Good Health and Well-being), SDG 6 (Clean Water and Sanitation), and SDG 10 (Reduced Inequalities), which are crucial for addressing the disease.

**Author summary:** Leprosy remains a serious public health issue in Brazil and other endemic countries, often tied to social and environmental inequalities. We conducted a comprehensive review of 44 studies published between 2015 and 2024 to understand the spatial distribution of leprosy in urban areas. We found that the disease doesn’t spread randomly; it consistently clusters in vulnerable, peripheral neighborhoods.

Our analysis confirms a strong link between leprosy and key socioeconomic factors like low income, poor education, and inadequate sanitation. The presence of new cases in children signals ongoing active transmission within these communities. The findings highlight that fighting leprosy requires more than just medical treatment. We must also address the underlying structural inequalities, aligning health efforts with broader social and urban policies, such as those outlined in the Sustainable Development Goals (SDGs). This approach will allow us to create more effective and targeted public health strategies.

## Introduction

Leprosy remains an important public health problem despite advances in control achieved over recent decades [1]. It is estimated that in 2022, more than 174,000 new cases were diagnosed worldwide, concentrated in endemic countries such as India, Brazil, and Indonesia [2]. Brazil ranks second globally in absolute number of cases, accounting for approximately 12% of global notifications, and continues to report high detection rates in certain regions, particularly in the North, Northeast, and Central-West [3].

Although global prevalence has declined since the implementation of multidrug therapy, leprosy continues to affect socially vulnerable populations, reflecting its close relationship with poverty, social inequality, and limited access to health services [4–6]. Beyond its clinical outcomes and disabling sequelae, the persistence of transmission clusters in urban areas represents an additional challenge for elimination programs [7,8].

Traditional epidemiological studies have contributed to leprosy surveillance; however, the spatial dimension of the disease remains underexplored in many regions. Spatial analysis enables the identification of high-risk areas, the evaluation of aggregation patterns, and a better understanding of the influence of socio-environmental determinants on transmission, thus becoming a strategic tool for health surveillance [7–10].

Accelerated urbanization, characterized by unplanned expansion, unequal access to basic infrastructure, and socio-spatial segregation, reinforces the need to understand how leprosy is distributed in urban territories [6,11,12]. In several Brazilian municipalities, high-risk clusters have been identified in peripheral neighborhoods with higher levels of vulnerability, confirming that the disease persists as a marker of social and environmental inequality [5,13].

In this context, it becomes essential to synthesize scientific production on the spatial distribution of leprosy in urban areas and its associated socio-environmental factors. Therefore, this systematic review aimed to analyze the literature published between 2015 and 2024, identifying recurrent spatial patterns, methodologies employed, and social and environmental determinants associated with leprosy, to support public policies and health surveillance strategies to address the disease.

## Methodology

This study is a systematic review of the literature, conducted in accordance with PRISMA 2020 guidelines [14], without meta-analysis. Its objective was to identify and synthesize studies that analyzed the spatial distribution of leprosy in urban areas and the associated socio-environmental factors. This review was registered in the International Prospective Register of Systematic Reviews (PROSPERO) under the number CRD420251129311.

### Search strategy

The bibliographic search was performed in PubMed/MEDLINE, Scopus, Web of Science, LILACS, and SciELO. In addition, a search of gray literature was conducted via Google Scholar, from which only articles published in peer-reviewed scientific journals and one relevant preprint were included. Thesis, dissertations, monographs, and institutional documents were excluded, as they did not meet the peer-review requirement.

Standardized descriptors (MeSH and DeCS) combined with Boolean operators (AND/OR) were used, such as:

- “Leprosy” OR “Hansen’s disease”
- “Spatial analysis” OR “Geographic Information Systems” OR “GIS”
- “Spatial distribution” OR “Spatial epidemiology”
- “Urban health” OR “Cities” OR “Urban areas”
- “Social determinants of health” OR “Socioeconomic factors”

Examples of applied combinations included: “Leprosy AND Spatial analysis”, “Hansen’s disease AND Geographic Information Systems”, and “Leprosy AND Urban health”. The time frame covered the years 2015 to 2024, and publications in Portuguese, English, and Spanish were included.

### Inclusion criteria

Studies were included if they:

- Investigated leprosy in an urban context (municipalities, neighborhoods, census tracts, or urban stratifications in broader analyses).
- Aimed to analyze the spatial distribution of leprosy and/or investigate associated socioeconomic, environmental, or demographic factors.
- Applied at least one spatial analysis technique, such as georeferencing and thematic mapping, Kernel Density Estimation (KDE), spatial autocorrelation (Global Moran’s I and Local Moran’s I/LISA), spatial/temporal scan statistics (SaTScan), spatial models (OLS, GWR, Spatial Lag/Error), Bayesian smoothing, or hotspot analysis.
- Were published in peer-reviewed journals or as relevant preprints.

### Exclusion criteria

Studies were excluded if they:

- Did not present an urban or intra-urban stratification in the results.
- Reported only epidemiological data without spatial analysis.
- Were narrative reviews or opinion pieces without quantitative analysis.
- Focused exclusively on rural, Indigenous, or prison populations without urban stratification.
- Were thesis, dissertations, monographs, or institutional documents not published in peer-reviewed journals.

### Research question: PECO framework

The guiding question of this review was “What are the spatial distribution patterns of leprosy in urban areas, and which environmental and socioeconomic factors are associated with its occurrence?”

The question was structured using the PECO framework:

- P (Population): individuals with leprosy living in urban areas.
- E (Exposure): socioeconomic, environmental, and demographic factors.
- C (Comparison): different population and territorial strata.
- O (Outcomes): spatial distribution of leprosy, cluster identification, and associated factors.

### Study selection

Screening was carried out in Rayyan software [15] in two stages:

1. Title and abstract screening to exclude studies clearly unrelated to the topic.
2. Full-text review for strict application of inclusion and exclusion criteria.

Selection was performed independently by two reviewers. Agreement between reviewers was assessed using the Kappa index, which yielded a value of 0.622, indicating substantial agreement according to Landis & Koch [16].

### Quality assessment of studies

The methodological quality of included studies was assessed using the Joanna Briggs Institute (JBI) Critical Appraisal Checklist for Analytical Cross-Sectional Studies [17], complemented by criteria specific to spatial analyses (geographic unit, spatial autocorrelation, geocoding, adjustment for multiple testing, and sensitivity analysis). Each study was classified as high, moderate, or low quality according to the proportion of criteria met. The full checklist is presented in Appendix 1.

### Data extraction and analysis

Data were extracted in a standardized spreadsheet, including title, authors, year, country/region, study design, geographic scope, socioeconomic and environmental variables analyzed, spatial analysis techniques, associations found, presence of clusters, high-risk populations, and main conclusions.

The analysis was descriptive and comparative, seeking to identify recurrent spatial patterns, key determinants, and contextual differences across countries, regions, and cities.

### Ethical aspects

As this study is a systematic literature review, without direct involvement of human participants or individual-level data, submission to a research ethics committee was not required.

In line with the Committee on Publication Ethics (COPE) guidelines on authorship and the use of AI tools, we acknowledge that generative AI tools specifically ChatGPT [18] and Gemini [19] were used to support specific tasks such as formatting references, organizing synthesis tables, and providing academic writing support (particularly grammar correction and improvements in textual cohesion and clarity).

All AI-generated content was critically reviewed by the authors to ensure accuracy, methodological consistency, and scientific adequacy. The use of AI was strictly supportive and did not replace the authors’ critical judgment or interpretative analysis.

## Results

The process of identification, screening, eligibility, and inclusion of studies is summarized in the PRISMA flow diagram (Fig. 1). In total, 314 records were initially retrieved from databases, of which 44 articles fully met the inclusion criteria and were analyzed in this review.

**Fig. 1.**
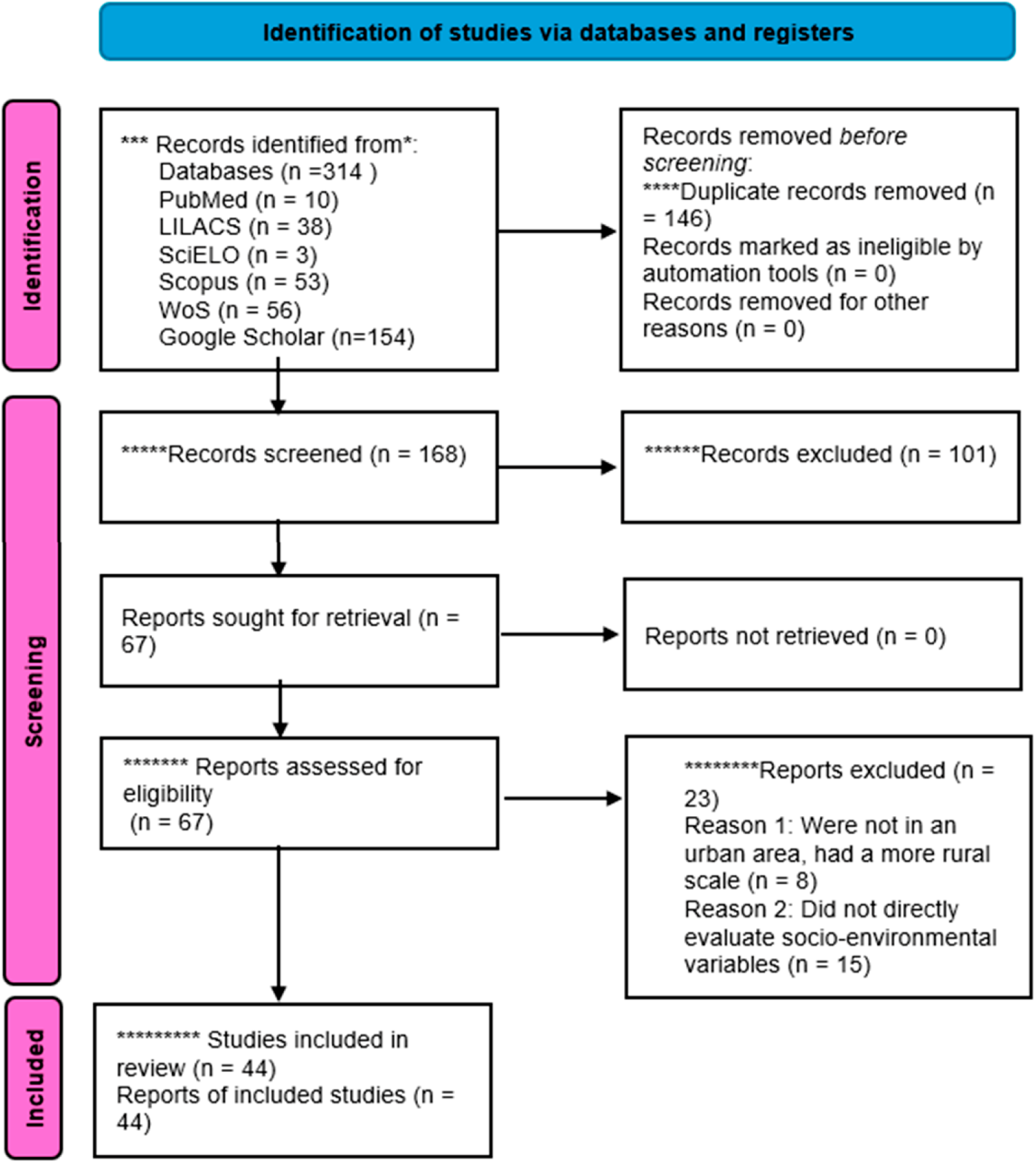
Flowchart of the study identification, screening, eligibility, and inclusion process, according to the PRISMA 2020 protocol.

### Methodological quality assessment

The methodological assessment of the 44 included studies, conducted with the adapted JBI checklist and additional spatial criteria (Appendix 1), showed that the vast majority were classified as high quality (39; 88.6%), while only 5 studies (11.4%) were rated as moderate quality. None were classified as low quality.

High-quality studies applied robust spatial modeling approaches (e.g., SaTScan, GWR, Bayesian models, spatial regressions) and thoroughly analyzed socio-environmental determinants. In contrast, moderate-quality studies presented limitations such as purely descriptive analyses, lack of confounder adjustment, or insufficient methodological validation.

### General characteristics of the included studies

The 44 studies were published between 2015 and 2024, covering diverse geographic contexts, with a predominance of Brazil (n = 38), followed by India, Indonesia, Bangladesh, Nigeria, and Madagascar.

Most studies adopted an ecological and cross-sectional design, using secondary data from health information systems (especially SINAN) and population censuses (Table 1).

**Table 1:** General characteristics of the 44 studies included in the systematic review, according to authors, year, study design, and spatial analysis techniques.

| Authors, Year | Study Design | Spatial Analysis Techniques |
| --- | --- | --- |
| Geice Silva Clemente, Delma Holanda de Almeida, 2024 [20] | Ecological, spatial and spatiotemporal analysis | Global Moran, LISA, spatiotemporal scanning (Poisson), Bayesian estimator |
| Edson J. C. Queiroz, Ingrid N. Rocha, et al., 2024 [12] | Cross-sectional, analytical, descriptive and quantitative | Georeferencing, QGIS 3.34, SPSS clustering, RR calculation |
| Meuthia Jasmine, Arief Wibowo, 2024 [21] | Quantitative, spatial analysis | Moran's I, LISA, Queen spatial weights |
| Amanda G. Carvalho, Carolina L. H. Dias, David J. Blok, Eliane Ignotti, João G. G. Luz, 2023 [9] | Ecological | SaTScan, OLS, SLM, SEM, GWR, Moran's I |
| Gutemberg H. Dias, José A. C. França, Filipe S. Peixoto, Caio A. M. Aires, 2023 [22] | Descriptive with spatial and epidemiological analysis | QGIS 3.16, ArcMap 10.8, heatmaps (Kernel) |
| Isabela C. Bueno, Daniele S. Lages, Francisco C. F. Lana, 2023 [23] | Ecological | Epidemiological risk index, Moran (global/local), Bayesian methods |
| José França, Caio Aires, Maurício Nobre, Sidnei Sakamoto, Gutemberg H. Dias, 2023 [24] | Ecological, descriptive and spatial | Geocoding, Voronoi, Kernel, proximity analysis |

| **Authors, Year** | **Study Design** | **Spatial Analysis Techniques** |
| --- | --- | --- |
| Vanessa M. S. Duarte, Lúbia M. G. Machado, Emerson S. Santos, Amílcar S. Damazo, 2023 [25] | Epidemiological observational, descriptive, ecological | Empirical Bayesian, spatial scanning (SatScan), GeoDa, ArcGIS |
| Freitas LRS, Duarte E.C., 2022 [26] | Retrospective ecological | Moran's I, Kernel, SaTScan |
| Romário R. Sousa, Hildeu F. Assunção, et al., 2023 [27] | Epidemiological, spatial and temporal | Kernel, geocoding, ArcGIS |
| Antônio C. V. Ramos, José F. Martoreli Jr., Thaís Z. Berra, et al., 2022 [28] | Ecological, spatial and spatiotemporal | Kernel, spatial and spatiotemporal scanning, SVTT |
| Emanuele R. Silva, Marcos Adami, Waltair M. M. Pereira, 2022 [29] | Ecological, temporal and spatial | Joinpoint, SaTScan, Poisson |
| Maria S. C. Linhares, Lígia R. F. S. Kerr, et al., 2022 [11] | Cross-sectional descriptive with spatial analysis | Georeferencing, Voronoi, NNI (nearest neighbors) |
| Ian C. A. Barros, Carliane C. M. Sousa, et al., 2024 [30] | Ecological | SaTScan, Joinpoint, LISA |
| A. T. Taal; D. J. Blok; A. Handito; et al., 2022 [31] | Observational ecological with spatial analysis | Moran's I, Kernel Density, hotspot analysis (QGIS) |
| Antônio Carlos V. Ramos, Jonas B. Alonso, et al., 2021 [32] | Ecological, descriptive with spatial analysis | Kernel, Moran's I, LISA |
| Caroline A. Bulstra, David J. Blok, et al., 2021 [33] | Retrospective observational (20 years) | Geospatial Poisson model, SaTScan, Empirical Bayesian |
| Fernanda C. Lopes, Giana G. S. Sousa, et al., 2021 [34] | Ecological | Geocoding (TerraView), SaTScan |
| Yasmin P. Azevedo, Vitor A. S. Bispo, et al., 2021 [35] | Descriptive epidemiological, secondary data | Google Earth Pro, QGIS 2.18.14, thematic maps |
| Gabriela C. Ribeiro, Josafá G. Barreto, Isabela C. Bueno, et al., 2021 [36] | Cross-sectional, descriptive and analytical | Georeferencing, spatial clusters, QGIS |
| Jaqueline R. Borba, Jéssica M. Penso-Campos, et al., 2021 [37] | Quantitative, descriptive and cross-sectional | Global Moran, GLM, TabWin, TerraView, ArcGIS, SPSS |
| Jucileide M. Alves, Roquenei P. Rodrigues, Monalisa C. S. Carvalho, 2021 [38] | Epidemiological, historical series | Average incidence, choropleth maps (QGIS) |
| Celivane C. Barbosa, Cristine V. Bonfim, et al., 2020 [39] | Ecological, spatial analysis | Empirical Bayesian, Moran (global/local), TerraView, QGIS |
| Aldenyessle R. Albuquerque, José V. M. Silva, et al., 2020 [40] | Mixed ecological | Moran's I, LISA, risk maps |
| Juliana B. Macedo, Daniela B. Macedo, et al., 2020 [41] | Ecological, analytical and exploratory | Global Moran/LISA, Moran maps |
| Carlos D. F. Souza, Carlos F. Luna, Mônica A. F. M. Magalhães, 2019 [5] | Mixed ecological | Joinpoint, SaTScan, Poisson, QGIS |
| Joilda S. Nery, Anna Ramond; et al., 2019 [42] | Ecological, spatial/temporal | Moran, SaTScan, LISA, Bayesian |
| Delma B. Sousa, Reinaldo Souza-Santos, et al., 2019 [43] | Ecological with individual analysis | Google Earth, Kernel, QGIS |
| Márcio B. Santos, Allan D. Santos, et al., 2019 [44] | Ecological and time series | Moran (Global/LISA), Kernel, Poisson regression |
| Flávia R. Ferreira, Luiz F. C. Nascimento, 2019 [45] | Ecological, exploratory | Moran, Kernel, Bayesian, TerraView |
| Carlos D. F. Souza, Carlos F. Luna, Mônica A. F. M. Magalhães, 2019 [46] | Ecological | Empirical Bayesian, Moran (global/local), spatial regressions |
| Ivaneliza S. de Assis, Marcos A. M. Arcoverde, et al., 2018 [47] | Ecological, spatial and temporal | Geocoding, SatScan, Moran, OLS, GWR, ArcGIS |
| Celivane C. Barbosa, Cristine V. Bonfim, et al., 2018 [48] | Ecological | Empirical Bayesian, Moran, LISA |
| C.D.F. de Souza, V.S. Rocha et al., 2019 [49] | Spatial/temporal ecological | SaTScan, TerraView, Poisson |
| Lúcia R. S. de Freitas, Elisabeth C. Duarte, Leila P. Garcia, 2017 [50] | Ecological with spatial analysis | SaTScan (Poisson), ArcGIS 9.2 |
| Ariella C. Luz, Victorugo G. A. Correia, et al., 2017 [51] | Descriptive, cross-sectional, retrospective | GPS, QGIS, Google Earth, thematic maps |
| Antônio C. V. Ramos, Mellina Yamamura, et al., 2017 [52] | Descriptive and ecological | Geocoding, SatScan, ArcGIS |
| Lorena D. Monteiro, Rosa M. S. Mota, et al., 2017 [4] | Ecological, secondary data | Negative binomial log-linear regression, Empirical Bayesian |
| Emanuele C. Chaves, Samara V. Costa, et al., 2017 [53] | Ecological, cross-sectional | Bivariate Moran, cluster analysis |
| Mônica Duarte-Cunha, Andréa S. Almeida, et al., 2016 [54] | Ecological | OLS, GWR, Moran, thematic maps |
| Veronique Suttels, Tom Lenaerts, 2016 [55] | Epidemiological and exploratory spatial | ArcGIS, Getis-Ord G, inverse distance |
| Mauricio E. S. Rangel, Ligia V. Barrozo, 2015 [56] | Ecological with spatial analysis | Crude and smoothed rates, TerraView, neighborhood matrix |
| L. D. Monteiro, F. R. Martins-Melo, et al., 2015 [13] | Ecological, spatial | Empirical Bayesian, Moran, maps |
| Mônica Duarte-Cunha, Geraldo M. da Cunha, Reinaldo Souza-Santos, 2015 [57] | Retrospective spatial ecological | Moran's I, LISA, Empirical Bayes, SaTScan |

### Geographic scope of analyses

The predominant spatial scale was intra-urban, with studies conducted in Brazilian municipalities such as Cuiabá (MT), Mossoró (RN), Sobral (CE), Imperatriz (MA), Feira de Santana (BA), Arapiraca (AL), and Paulo Afonso (BA).

Some studies applied state- or regional-level analyses when urban stratification was available (e.g., Bahia, Pará, Tocantins, Piauí, Rondônia, and Minas Gerais). In Asian and African countries, the most frequent scale was the district level, often with urban detail.

### Socioeconomic and environmental variables analyzed, and spatial analysis techniques used

Among the most frequently investigated socioeconomic variables, income was assessed in 16 studies (36.4%) and education in 13 (29.5%). Other recurrent determinants included poverty (9 studies; 20.5%), race/skin color (11; 25.0%), the Gini Index (6; 13.6%), and the Municipal Human Development Index (HDI-M) (5; 11.4%). Only three studies (6.8%) analyzed social inequality in a generic manner, and a single study (2.3%) included household density as the main variable.

In the environmental domain, the most frequently reported variables were basic sanitation and access to piped water (5 studies each; 11.4%), followed by rudimentary pit latrines and waste collection (3 studies each; 6.8%). Deforestation was analyzed in two studies (4.5%), and unplanned urban expansion in only one (2.3%).

The methodologies applied varied widely across studies. The most recurrent were spatial autocorrelation tests (Global/Local Moran’s I), employed in 21 studies (47.7%). Other frequently used techniques included SaTScan (14; 31.8%) and Bayesian estimators (12; 27.3%). Additional methods such as LISA, Kernel density estimation, and GIS software (QGIS, ArcGIS, TerraView) appeared in approximately one quarter of the studies. In contrast, more advanced approaches such as Geographically Weighted Regression (GWR) and spatial regression models (OLS, Spatial Lag, Spatial Error) were rare, occurring in only 3 studies (6.8%). These findings reinforce that socioeconomic deprivation was the most consistent determinant, while environmental factors were addressed in a more limited way (Table 2).

**Table 2:** Distribution of socioeconomic and environmental variables and spatial analysis techniques used in the studies included in the review (n = 44)

| Category | Item | No. of studies | % |
| --- | --- | --- | --- |
| Socioeconomic | Income | 16 | 36.4 |
| Socioeconomic | Education | 13 | 29.5 |
| Socioeconomic | Poverty | 9 | 20.5 |
| Socioeconomic | Race/skin color | 11 | 25.0 |
| Socioeconomic | Gini Index | 6 | 13.6 |
| Socioeconomic | HDI-M | 5 | 11.4 |
| Socioeconomic | Social inequality | 3 | 6.8 |
| Socioeconomic | Household density | 1 | 2.3 |
| Environmental | Basic sanitation | 5 | 11.4 |
| Environmental | Piped water | 5 | 11.4 |
| Environmental | Pit latrines | 3 | 6.8 |
| Environmental | Waste collection | 3 | 6.8 |
| Environmental | Deforestation | 2 | 4.5 |
| Environmental | Unplanned urban expansion | 1 | 2.3 |
| Technique | Moran (Global/Local) | 21 | 47.7 |
| Technique | SaTScan | 14 | 31.8 |
| Technique | Bayesian methods | 12 | 27.3 |
| Technique | LISA | 10 | 22.7 |
| Technique | Kernel density | 10 | 22.7 |
| Technique | QGIS | 10 | 22.7 |
| Technique | ArcGIS | 7 | 15.9 |
| Technique | TerraView | 6 | 13.6 |
| Technique | GWR | 3 | 6.8 |
| Technique | OLS | 3 | 6.8 |

### Clusters and High-Risk Populations

The studies identified persistent high-risk clusters in urban peripheral areas marked by greater socioeconomic vulnerability.

- In Brazil, clusters were more frequent in the North and Northeast regions, especially in peripheral neighborhoods and areas of urban expansion.
- In Asia, districts in India and Bangladesh showed high endemic clusters in densely populated and impoverished areas.
- The populations most affected included children under 15 years of age (indicating active transmission), adult men of working age, residents of areas with inadequate sanitation, and families living under conditions of structural poverty.

## Discussion

This systematic review demonstrated that spatial analysis is an essential tool to understand the heterogeneous distribution of leprosy in urban areas, allowing the identification of high-risk clusters, persistent transmission patterns, and associated socio-environmental factors. Studies conducted in different contexts confirm that the endemic persists unevenly, concentrated in pockets of vulnerability, particularly in the North and Northeast regions of Brazil [3,5,13].

### Socioeconomic determinants

Among the social determinants, low income and low education were the factors most frequently associated with leprosy occurrence. In Tocantins, the spatial analysis of more than 14,000 cases revealed a strong association with social inequality indicators and clusters in highly vulnerable areas [13]. Similar results were reported in Bahia and Minas Gerais, where high-risk clusters coincided with neighborhoods of low schooling and reduced income [5,23]. Racial inequality also emerged as an important determinant, with greater risk observed among mixed-race, Black, and Indigenous populations [11,12]. These findings align with international evidence, such as in Bangladesh, where leprosy hotspots persisted in areas of extreme poverty and high population density [33].

### Environmental determinants

Although less explored, environmental variables showed relevance in the studies. In Sobral and Mossoró, factors such as poor sanitation, rudimentary pit latrines, and lack of waste collection were associated with leprosy clusters [11,58]. In Imperatriz, Maranhão, unplanned urban expansion and the proximity of highways and deforested areas were also identified as environmental conditions influencing the disease [34].

In Pará, the Social Deprivation Index (SDI) was developed using 2010 Census variables related to sanitary conditions (absence of toilets, lack of waste collection, lack of piped water) and socioeconomic conditions (low household income and low educational level of household heads). This index, constructed through factor analysis and classified into three categories (good, fair, poor), showed a positive spatial correlation with leprosy, indicating that municipalities with greater social deprivation, especially in the Marajó and Araguaia regions, had higher detection rates [53]. These findings reinforce that the combination of socioeconomic and environmental determinants increases the risk of transmission but also highlight a methodological gap, as environmental variables are still rarely incorporated.

### Spatial analysis techniques

The methodologies applied demonstrated progress but also limitations. Moran’s I and its variants (LISA) were the most frequently used techniques, identifying significant spatial dependence in different contexts [13,21]. SaTScan was widely employed to detect spatio- temporal clusters in hyperendemic states and in nationwide analyses [3,46].

Bayesian approaches, such as the local empirical estimator, were applied in Maranhão and Pernambuco and proved effective in reducing random fluctuations and identifying priority areas [48,56]. More robust techniques, such as Geographically Weighted Regression (GWR) and spatial regression models (OLS, Spatial Lag, Spatial Error), were rarely explored but performed well in studies from Cuiabá and the tri-border region [9,47]. These findings highlight the need to expand the use of advanced methods to better capture intra-urban heterogeneity.

### Clusters and high-risk populations

The occurrence of leprosy in children under 15 years was reported in several contexts and is considered an important marker of active transmission and failures in early detection, as it indicates recent community infection [10,11]. At the same time, the predominance of the disease among adult men of working age suggests the influence of occupational factors, housing conditions, and unequal access to health services on the persistence of endemicity [35,38]. These findings reveal that both children and adults of working age represent priority groups for active surveillance and prevention strategies, requiring intersectoral actions integrating health, education, labor, and social policies.

### International Comparison

The international analysis showed that the determinants of leprosy in Asian and African contexts resemble those found in Brazil. In Madagascar, leprosy was concentrated in urban districts with high population density and poor living conditions [55]. In Indonesia, spatial analyses identified hotspots in urban regions, providing evidence for prophylactic strategies among contacts [21,31].

In Bangladesh, despite the global decline in cases, high-risk clusters persisted in dense and poor areas [33]. These findings converge with the Brazilian literature, reinforcing the importance of policies targeting social and urban inequalities.

### Public health implications

The synthesis of findings reinforces that leprosy remains a marker of social and environmental inequalities. The identification of urban clusters can guide territorially oriented surveillance actions and strengthen primary health care, particularly through the Family Health Strategy (ESF).

Recent experiences, such as the use of rifampicin chemoprophylaxis (SDR-PEP) in Alta Floresta [59] demonstrate the potential of integrating spatial analysis with community-based interventions. Furthermore, the association between Bolsa Família Program (Family Allowance Program, Brazilian Government) coverage and reduced risk in hyperendemic municipalities highlights the impact of social policies on health outcomes [4].

This review’s findings underscore that the persistence of leprosy is a direct reflection of structural inequalities and poor living conditions, aligning with the objectives of the Sustainable Development Goals (SDGs). Specifically, the strong association found between the disease and low income, low education, and poor sanitation directly addresses the targets of SDG 1 (No Poverty), SDG 3 (Good Health and Well-being), SDG 4 (Quality Education), SDG 6 (Clean Water and Sanitation), and SDG 10 (Reduced Inequalities). The use of spatial analysis as a surveillance tool, as highlighted in this study, represents a strategic approach to achieving these goals by enabling more effective, targeted public health interventions.

### Gaps and recommendations

Despite methodological advances, important gaps remain. Environmental variables are underutilized and rarely incorporated into more complex analyses. The use of advanced spatial models is still limited, even in urban contexts where spatial heterogeneity is striking. There is also a lack of longitudinal studies integrating multiple determinants, which would be essential to understanding mechanisms of endemic persistence.

It is recommended that future research expand the use of robust spatial techniques, incorporate environmental variables, and evaluate intersectoral interventions integrating health, urban planning, and social assistance.

This systematic review shows that although leprosy has presented a downward trend in detection rates in Brazil and other endemic countries, the persistence of clusters in vulnerable urban areas, the occurrence in children under 15, and the association with social and environmental determinants demonstrate that elimination of the disease is still distant. The findings reinforce the need to integrate health and social policies, strengthen territorially oriented surveillance, and invest in innovative methodological approaches to understand and address the complexity of leprosy in cities.

### Final considerations

Leprosy in urban areas remains closely linked to social and environmental determinants, reflecting structural inequalities that persist in Brazil and other endemic countries. Spatial analysis has been consolidated as an essential tool for epidemiological surveillance, enabling the identification of persistent clusters, the most vulnerable populations, and priority areas for intervention.

The findings show that although there have been advances in diagnosis and treatment, disease elimination is still distant, mainly due to ongoing active transmission in children under 15 and the concentration of cases in poverty-stricken territories. In this sense, the integration of public health policies with broader social actions is critical to reducing the inequalities sustaining endemicity.

Future research should incorporate more robust spatial methodologies, expand the use of environmental variables, and evaluate intersectoral interventions, strengthening the articulation between health, urban planning, and social assistance. This approach will allow progress in understanding the urban dynamics of leprosy and contribute to the development of more effective strategies for disease control and elimination.

## Data Availability

All data are fully available, without restriction.

## Other Information

### Support

This research received no specific grant from any funding agency in the public, commercial, or not-for-profit sectors.

### Competing Interests

The authors declare that they have no competing interests.

### Availability of Data and Materials

All relevant data are within the paper and its Supporting Information files.

## Funding

The authors received no specific funding for this work.

## Competing interests

The authors have declared that no competing interests exist.

## Supporting Information

### Appendix 1: Joanna Briggs Institute (JBI) Critical Appraisal Checklist for Analytical Cross-Sectional Studies

Checklist used for the methodological appraisal of studies included in this systematic review, following the guidelines of the Joanna Briggs Institute (JBI).

1. Were clear criteria for inclusion in the sample included?
2. Were the study subjects and context described in detail?
3. Was the exposure measured in a valid and reliable manner?
4. Were the criteria used to measure the condition/disease objective, standardized, and valid?
5. Were confounding factors identified?
6. Were strategies to deal with confounding factors described?
7. Were the outcomes measured in a valid and reliable way?
8. Was an appropriate statistical analysis used?

**Source:** Joanna Briggs Institute. JBI Critical Appraisal Tools. JBI, 2020. Available from: https://jbi.global/critical-appraisal-tools.

## Notes

### Competing Interest Statement

The authors have declared no competing interest.

### Funding Statement

The author(s) received no specific funding for this work.

